# HARMONY (HARM reduction for Opiates, Nicotine and You) Trial: Protocol of a Randomised Controlled Trial of the Effectiveness of Vaporised Nicotine Products for Tobacco Smoking Cessation amongst NSW Opiate Agonist Treatment Clients

**DOI:** 10.1101/2024.06.21.24309014

**Authors:** B Bonevski, M Jackson, E Austin, N Lintzeris, N Ezard, C Gartner, C Oldmeadow, P Haber, R Hallinan, C Rodgers, T Ho, M Nean, M Harrod, A Dunlop

## Abstract

**Introduction:** Tobacco smoking is a major cause of preventable disease in Australia. Individuals receiving opiate agonist treatment (OAT) are a group who experience high tobacco-related morbidity and mortality rates. Despite reporting a desire to stop, relapse rates in OAT clients are high and cessation attempts supported by pharmacotherapy are less effective than in general populations. New and innovative ways of addressing smoking amongst this group are needed.

Vaporised nicotine products (VNPs), or e-cigarettes, may reduce a person’s exposure to toxicants and carcinogens when compared to tobacco cigarettes. High quality evidence indicates that VNPs can increase rates of smoking cessation compared to nicotine replacement therapy. Pilot results of VNPs as a smoking cessation aid in OAT clients suggests their use is feasible and acceptable but effectiveness in this group has not been explored.

This protocol details the rationale and methodology for a randomised controlled trial to examine the effectiveness of VNPs for tobacco smoking cessation amongst OAT clients in New South Wales, Australia.

**Methods and Analysis:** This will be a randomised single-blinded parallel group trial comparing 12-weeks of 12mg/mL vaporised nicotine to best-practice NRT. Participants must be 18 years or older, accessing opiate treatment at a participating health site, and a current daily tobacco smoker seeking to quit or reduce their smoking.

The primary outcome will be self-reported 7-day point prevalence abstinence from tobacco after 12-weeks of treatment. Secondary outcomes include biochemically verified abstinence, self-reported 30-day abstinence, number of cigarettes smoked each day, craving and withdrawal symptoms, and VNP safety. Between-group comparisons will be conducted at end of treatment, and at 12-weeks post-treatment.

**Discussion:** This study examines new ways of reducing tobacco related harm in individuals receiving OAT. Outcomes may be enhanced by leveraging participants interactions with health care provides who can facilitate the required support. Study findings have the potential to significantly impact tobacco smoking prevalence in priority populations.

**Ethics and Dissemination:** Protocol approval was granted by Hunter New England Human Research Ethics Committee (Reference 2020/ETH01866). Findings will be disseminated via academic conferences, peer-reviewed publications and social media.

**Registration:** The study was registered in the Australian New Zealand Clinical Trials Registry (Reference ACTRN12621000148875).

## INTRODUCTION

Smoking continues to be the main cause of preventable disease in Australia linked to cancer, respiratory disease and cardiovascular disease.(1) Although smoking rates have decreased in the general population in Australia (13%), they continue to be disproportionately high amongst lower socioeconomic status groups.(2, 3) An approach that combines population-based tobacco control (e.g. mass media) with targeted interventions for high prevalence groups is recognised as likely to yield the greatest benefits.(2, 4, 5) For the purposes of this protocol, the word “smoking” is defined as tobacco smoking. All references to smoking and smoking cessation herein refers to tobacco cigarette smoking and not, for example, cannabis or e-cigarette smoking.

### Rationale for performing the study

Tobacco smoking is highly prevalent amongst opiate agonist treatment clients. The highest rates of smoking amongst people with substance use disorders are seen in those on opiate agonist treatment (OAT): as high as 73% to 94% smoking prevalence. As a result, people seeking treatment for opiate dependence experience a greater tobacco-related burden of illness. (6) In a Swedish study, (7) OAT clients were observed to have an excess mortality rate of 18.3 times the standardized mortality rate (SMR). Causes of death were largely due to tobacco related diseases: neoplasms were 2.2 times the SMR, circulatory causes were 3.9 times the SMR, and respiratory causes 2.5 times the SMR. Data from 140 clients on methadone over the age of 50 in one US study (8) demonstrated a high prevalence of cardiovascular and chronic lung disease. Current smokers comprised 87.1% of interviewed subjects. Among these smokers, 17.9% had an active heart condition and 22.1% had chronic lung disease. Tobacco cessation profoundly improves people’s lives reducing risk of disease, improving quality of life, reducing depression, anxiety and financial stress. (9)

Smokers receiving OAT are interested in quitting. Clients in OAT are greatly interested in quitting smoking, especially if provided in their treatment clinic. (10) Addressing smoking during treatment does not jeopardise other outcomes such as retention in opiate treatment. One review of 24 studies has found that smoking cessation does not negatively affect achievement of other drug treatment goals and can enhance them. (11)

However, relapse to smoking is common. While 85% of quit attempts using smoking cessation support fail in the long term (e.g. 12 months) among smokers in the general population, this rate is even higher amongst smokers with opiate use. While smoking cessation appears possible in OAT smokers, few interventions have demonstrated efficacy in this population of tobacco users. (12) Most interventions decreased cigarette use, but sustained abstinence rates remained low. High relapse rates in this population may be due to factors related to addiction (e.g. nicotine enhancing the opioid withdrawal reductions due to OAT), lack of cessation support, and the high levels of smoking in their social network. (13) All of the randomized trials of smoking cessation pharmacotherapies tested in persons with opioid use disorder, including NRT, (14) bupropion, (15) and varenicline (16) have produced far lower quit rates than those reported in trials of nondrug users. New ways of thinking about how to address smoking amongst these populations are needed.

### Vaporised nicotine products for smoking cessation

VNPs are a broad range of battery-powered devices that deliver an aerosol of propylene glycol and/or glycerine, nicotine and flavours. (17) Unlike combustible tobacco cigarettes, VNPs deliver nicotine in an inhalable form without burning tobacco. Like pharmaceutical NRT, the provision of nicotine in VNP aerosol reduces cravings and withdrawal symptoms. With practice VNP users can obtain similar blood nicotine levels to cigarette smokers. (18, 19) VNP aerosol may still contain some toxicants, especially if the liquid is overheated, but these are much fewer than in cigarette smoke and those that are present are at much lower concentrations.(20) Weighing up all the available safety data, a 2018 committee report by the US National Academies of Science, Engineering and Medicine concluded that substituting VNPs for combustible tobacco cigarettes conclusively reduces a person’s exposure to many toxicants and carcinogens present in combustible tobacco cigarettes and may result in reduced adverse health outcomes. (21)

Many smokers use VNPs to quit smoking tobacco cigarettes. (22) Large scale population surveys in the US (n=161,054) (23) and UK (n= 170,490) (24) suggest that VNP use is significantly associated with smoking cessation. A Cochrane review of RCTs found that VNPs are at least as effective as other forms of NRT. (25) The authors noted however the limited number of studies (2 RCTs with combined n=662) and recommended more RCTs be conducted. The most recent large RCT published in NEJM 2019 with clients of UK Stop Smoking Services found that VNPs almost doubled the odds of long-term cessation compared with NRT (18% vs 10%; RR=1.83; 95% CI, 1.30 to 2.58; P<0.001). (26) Research to date with VNPs for smoking cessation have been mainly conducted with smokers in the general population. A small (n=12) single group trial in the USA among people on OAT found reductions in cigarettes smoked per day over a 6 week, VNP-use study period. (27) There are no other trials of VNPs for smoking cessation amongst OAT clients published or registered.

### Smoking cessation in people receiving opioid agonist treatment

A pilot study of 50 OAT clients in Newcastle, NSW showed that enrolment into a clinical trial of VNPs for smoking cessation was feasible (90% recruitment rate and 84% retention rate at follow-up); acceptable (8/10 satisfaction rating; 9/10 helpful for quitting); and potentially effective (continuous abstinence rate 33% for gradual reduction methods and 41% for abrupt cessation method at follow-up). Adherence to the use of the VNPs was high (81%, with average use 9.4 times per day at follow-up). (28)

## OBJECTIVES

### Primary Objective

The primary aim of the study is to examine the effectiveness of treatment with VNP compared to best practice NRT on self-reported 7-day point prevalence abstinence from tobacco smoking at the end of 12 weeks of nicotine treatment in OAT clients.

### Secondary Objectives

- To compare the effectiveness of treatment with VNP compared to best practice NRT on biochemically verified self-reported 7-day point prevalence abstinence from tobacco smoking at the end of 12 weeks of nicotine treatment in OAT clients.
- To compare self-reported 30-day continuous abstinence from tobacco smoking for the VNP treatment group relative to the best practice NRT group at 3 months post-treatment.
- To compare self-reported 7-day point prevalence abstinence from tobacco smoking for the VNP treatment group relative to the best practice NRT group at 3 months post-treatment.
- To examine the safety of liquid nicotine delivered via VNPs in the study population, relative to best practice NRT.
- To determine if treatment with VNP relative to best practice NRT reduces the number of cigarettes smoked per day by OAT clients prior to treatment by 50% (or more) at end of treatment and 3 months post-treatment.
- To compare nicotine craving and withdrawal symptoms for participants in the two treatment groups (VNP vs best practice NRT) at end of treatment and 3 months post-treatment.
- To compare tobacco relapse episodes for participants in the two treatment groups (VNP vs best practice NRT) at end of treatment and 3 months post-treatment.
- To compare nicotine treatment adherence for participants in the two treatment groups (VNP vs best practice NRT) at end of treatment and 3 months post-treatment.
- Compare other drug use in the two groups (including cannabis use, amphetamine, non-prescribed opioids) at end of treatment and 3 months post-treatment.
- To compare study retention for participants in the two treatment groups (VNP vs best practice NRT) at end of treatment and 3 months post-treatment.
- A within trial analysis to compare the cost and consequence of treatments for participants in the two arms (VNP vs best practice NRT).

## METHODS

### Study Design

This is a randomised single blinded parallel group trial comparing a 12-week course of liquid nicotine delivered via VNPs to best practice NRT in a 1:1 ratio of study groups.

The study population are adults receiving OAT with nicotine dependence.

### Duration for Participants

Total duration of the study participation for each participant will be approximately 26 weeks; this consists of a screening period of up to 2 weeks, a treatment period of 12 weeks and a follow up period of 12 weeks.

### Participating Sites

This study is being conducted by drug and alcohol services in six New South Wales (NSW) Local Health Districts.

Sites may commence recruitment once all required trial documentation is in place and a site initiation (visit or teleconference) has taken place.

## PARTICIPANT SELECTION

### Inclusion Criteria

- Provide written, informed consent to participate in the study.
- Aged 18 years or older.
- Be accessing opioid agonist treatment from a participating service.
- Current daily tobacco smokers on self-report.
- Want to quit or cut down their smoking.
- Be willing and able to comply with requirements of study (including having access to a phone).

### Exclusion Criteria

- Currently breastfeeding or pregnant, or of childbearing potential and planning/trying to fall pregnant during the study period.
- Current, severe medical disorder assessed by study medical officer (such as but not limited to, unstable cardiovascular/peripheral vascular disease, poorly controlled hypertension)
- Current, severe or unstable psychiatric disorder assessed by study medical officer (such as but not limited to, acute psychosis, severe anxiety and/or mood disorder, intent to harm self or others).
- Current enrolment in a clinical trial involving any investigational drug 2
- Regular use (more than one day per week) of Vaporised Nicotine Product (or e-Cigarette) containing nicotine in the last 30 days.
- Not available for follow-up (e.g. likely imprisonment or transfer out of service to another service that is not a trial recruitment site).

## RECRUITMENT & RETENTION

### Referral

The recruitment strategy for this study will incorporate approved advertisements being displayed in participating services OAT waiting rooms and other suitable locations. Due to recent changes in OAT service delivery, advertisements may also be sent via text or email to clinic clients that are no longer visiting clinics daily. Interested clients may contact the researchers directly using contact details contained in the advertisements.

Introductions to the trial shall also be made by the clinicians at each site during appointments or conversations. Clinicians will be asked to identify clients who smoke and enquire about their desire to stop and their interest in participating in a smoking cessation treatment using the script provided. The contact details of any interested clients will be provided to the research team for pre-screening.

### Pre-Screening

All interested clients shall be considered for pre-screening. A RO/RN will complete a pre-screening questionnaire with the client either in person or by telephone. Potential eligible persons will be invited to attend the clinic in person or via Telehealth for an informed consent discussion and a formal screening assessment.

### Screening

De-identified information relating to each patient screened shall be recorded regardless of whether or not the individual is eligible. If the patient is ineligible, the reason will be recorded.

Screening will continue until the target population is achieved.

De-identified Screening Logs for this study will be generated electronically through the REDCap database by the Project Manager.

### Informed Consent

Patients will be provided with the current Human Research Ethics Committee (HREC) approved participant information sheet for their consideration.

The investigator, or a person designated by the investigator, will fully inform the client of all pertinent aspects of the trial. The client shall be given ample time and opportunity to inquire about details of the trial and to decide whether or not to participate in the trial. All questions about the trial will be answered to the satisfaction of the client.

Prior to a client’s participation in the trial, the informed consent form will be signed and personally dated by the client and by the Investigator (or delegate) who conducted the informed consent discussion. During this discussion, clients will be informed about the voluntary nature of the research, its content and what it involves for them. Clients will have the opportunity to ask questions before providing consent.

If clients cannot be physically present for the interview, then informed consent discussion may be completed over the phone or by teleconference, with electronic consent obtained using the REDCap eConsent Framework. This provides standardised tools to obtain consent and store consent documentation within the study database. In this case, clients will be sent the Patient Information Statement and Consent Form prior to discussion with the Investigator (or delegate). Discussions will be identical to those completed in person.

If a client is unable to read or has difficulty reading, the research member will read the information sheet to them and/or explain it in plain language, asking questions to ensure that its contents are understood.

### Eligibility Screening Procedures (Baseline)

Medical and eligibility screening will occur after informed consent has been obtained. The Screening Visit will include standard medical screening procedures as outlined in the following sections.

#### Screen Failures

If an individual consents to participate in the trial but are deemed ineligible prior to randomisation, they will be defined as a ‘screening failure’. Individuals identified as screening failures may be assessed for eligibility at a later time if their situation changes. Any screening failures are to be recorded on the screening log including the reason for screen fail. Any baseline measures that were completed at initial screening will need to be re-administered if the participant is rescreened more than 7 days later.

### Retention

Ensuring retention of participants for the End of Treatment assessment at Week 12 is vital to ensure the power and internal validity of the study (29). Participant attrition is a common problem in studies with people with substance use disorders. The following strategies will be used to assist treatment and study retention.

#### Phone contact

- Contact to be attempted by RO/RN during work hours.
- RO/RN to ask the participant if they have a preferred day and time to call.
- Obtain organisational approval to locate and access participant information.
- Include in the consent form a request to access medical records for personal details and emergency contact information.
- Ask the participant about other services that they are engaged with (e.g. community pharmacies, GPs, other social services) and if they would allow access to speak with the individuals working at these services to receive their updated details.

#### Educate and motivate

- During recruitment phase and during follow up visits, RO/RN is to explain to the participant the reasons for conducting the follow-up, when they can be expected to be followed up, the kinds of information that will be collected and how the information will be used.
- Inform the participant that if they relapse to smoking, we would still like to speak to them at Follow-up and at the End of Treatment.

#### Request information of contacts at baseline

- Check these contacts again and if they would like to add any additional contacts at Week 4, 8 & Week 12 visits.
- Must have a minimum of one alternate contact (e.g. significant other, another person in their household, worker from another service).

#### Multiple reminders

- Have flexible appointment times and call or send SMS reminders prior to all research appointments.

### Reimbursement

Participants will receive the following reimbursements using retail gift vouchers. These reimbursements are to cover time and expenses associated with study participation and are not deemed coercive. They are typical of amounts offered in studies of a similar nature.

- $50 gift card at completion of Week 12 interview.
- $50 gift card at completion of Week 24 interview.
- $50 gift card for participants that complete a CO breath test after reporting 7-day abstinence from tobacco at Week 12. These are to be provided on completion of the test and do not depend on test results.

## TREATMENT CONDITION ALLOCATIONS

### Condition 1: VNP

Participants randomised to Condition 1 shall be supplied with a VNP (Innokin Endura T18-II starter kit) and 12 weeks of prescribed liquid nicotine.

A one-week supply of NRT transdermal patches shall also be provided to participants randomised to Condition 1 for use whilst they are learning how to use the VNP effectively.

### Condition 2: NRT

Participants randomised to Condition 2 shall be supplied with a combination of NRT transdermal patches and oral forms (inhalators, gum, lozenges, mouth spray) to use throughout the 12-week intervention period.

### Adherence

Guidelines to improve adherence as well as information on nicotine product use and safety will be provided during the baseline interview. This will include:

- Importance of following study guidelines for adherence to the use of the VNP and/or NRT.
- A discussion on the length of time to use the VNP and/or NRT.
- Training on how to use the VNP and/or NRT correctly and safely.
- Training on how to access the New Zealand Ministry of Health Quit website https://vapingfacts.health.nz/ and encouragement to visit the site as an online resource throughout the trial.
- Importance of contacting the clinic or research team if experiencing problems possibly related to study product.

At each subsequent interview during the 12-week treatment period, participants will be asked about any problems they are having using the VNP and/or NRT. A brief discussion on reasons for problematic use and simple strategies for enhancing adherence, e.g. re-training on use of products will be undertaken. Participants will have an opportunity to ask questions and key messages from the initial session will be reviewed as needed.

### Randomisation

Participants will be randomised in a 1:1 ratio between groups using variable block randomisation stratified by treatment site and then by cannabis use (no use versus use, recorded by baseline ATOP). A computer-generated randomisation schedule will be developed by an independent statistician and uploaded to the REDCap study database.

Allocation concealment will be ensured by applying rules and user rights within the REDCap database. The randomisation allocation code will not be able to be generated until after the patient has been recruited have been completed.

Randomisation will be performed by the RO/RNs within the REDCap database according to the instructions included in the HARMONY Manual of Operations.

In the instance that two or more people from the same household wish to participate in the study at the same time, the first participant will be randomised as normal, the second will not be randomised but allocated the same treatment. This person will complete the study however their data will not be used in the main analysis.

### Blinding of outcome data assessors

To ensure blinding of the outcome assessments at Week 12 and 24, research interviews will be carried out by the Hunter New England Population Health Computer Assisted Telephone Interviewing (CATI) team who are independent of the participant and their trial related assessments.

#### CATI Team phone call schedule

The CATI team will contact participants to complete the week 12 and week 24 follow-up interviews using contact information provided by the study team. They will attempt to call 10 times over a 3-week period (-7 / +14 days from week 12 survey due date or -7 + 28 days from week 24 survey due date).

After contact is made for the Week 12 research interview, the CATI Team will check that the participant consents to being contacted again at Week 24.

Approved messages will be sent to each participant:

- SMS: 1-3 days prior to the survey due date.
- SMS 2: Following call attempt 5 (even if they have had contact)
- Voicemail: Leave message on call 2, and then again later if needed (e.g. Call 7).

In the event that the call schedule is exhausted, and the participant has not completed the week 12 or 24 follow-up interviews, the site research team member may contact the participant to complete the interview. Contact may be attempted for a period of 2 weeks after the final CATI call attempt.

## VISITS AND PROCEDURES

### Consent, Medical Screening and Baseline

The Investigator (or delegate) will conduct the following procedures during the screening period. This may occur in one visit or over multiple visits, depending on staff/client preferences:

- Informed consent (must be obtained prior to any other procedures being carried out)
- Clinical assessment (vital signs, weight)
- Medical history
- Substance use history, including any current use of VNPs
- Demographics
- Adapted Heaviness of Smoking Index
- Concomitant medications.

To facilitate follow-up, a Contact Information Form, including participant’s personal contact details and those of alternate contacts (e.g. family members, partners, friends, or services) will be completed.

### Treatment

Contact will be made with participants at Weeks 1, 4, 8 and 12 during treatment. These will occur either in person, over the phone or by teleconference and will be used to conduct research assessments, assess safety and provide nicotine products and training (where appropriate).

All attempts must be made to carry out research assessment interviews for the duration of the study regardless of whether the participant misses a visit or remains on treatment.

#### Baseline Visit (Week 1)

The Research Officer or Nurse will conduct the following procedures:

Adverse event review (since signing of consent form):

- Concomitant medication review.
- Provide the participant with nicotine product training, written information and wallet card.
- Set a quit date for the next two weeks.
- Provide written safety information about EVALI.
- Provide a letter to the participant for their GP or health care provider acknowledging their participation in the study, provided that such a practitioner can be identified for the participant.

Initial research assessment:

- Smoking and quitting history (including NRT use)
- Quit intentions / motivation / difficulty / self-efficacy
- Nicotine product usefulness
- VNPs – familiarity and use
- Heaviness of Smoking Index (HSI)
- Craving, Frequency and Strength
- Minnesota Tobacco Withdrawal Scale
- Amount spent on tobacco in the last week
- Psychological distress - Patient Health Questionnaire (PHQ-4)
- Substance use & wellbeing (ATOP)
- Current OAT treatment (medication, route of administration, dose, length of current treatment episode).

The Investigator (or delegate) will provide the following study medication and instruction on its use, as per the participant’s randomly assigned treatment. In instances where baseline procedures have been completed remotely, medication may be delivered to a verified address or pick up point (e.g. dosing pharmacy):

#### Condition 1: VNP

- Device loaded with one bottle of 12mg/ml e-liquid
- An additional seven (7) bottles of 12mg/ml e-liquid
- A brief information session on how to use the VNP
- A VNP information pack including safe storage of e-liquid nicotine and disposal
- 1-week supply of nicotine patches
- Training on the use of NRT patches
- Ensure that participant is comfortable with using the VNP and nicotine patch before leaving.

A prescription for the 12-week supply of nicotine e-liquid will be written and filled. The prescription will be kept as part of the participant’s confidential and secure study documentation. Participants will receive an initial 4-week supply of nicotine e-liquid, with the remainder supplied at or after Week 4 and 8 interviews.

In addition to pharmacy records, research staff will maintain a log of study products dispensed by the pharmacy when distributing to participants including recoding batch numbers on the Case Report Forms (CRFs).

#### Condition 2: NRT

- 28 x patches 21mg
- 2 x 20 pack of Inhalator 15mg
- 3 x canisters oral spray 1mg
- Training on the use of the NRT
- An NRT information pack including safe storage and disposal of used NRT products
- Where possible, ensure the participant leaves wearing an NRT patch and with an oral dose ready to use.

Participants will receive the initial 4-week supply of NRT as above (or equivalent). The remainder will be provided to the participant at or after the Week 4 and 8 interviews. The research team will supply all NRT provided as part of this study. No scripts or additional labelling will be required for dispensing to participants and NRT dispensed will be recorded in CRFs.

#### Monthly Visits (Weeks 4 and 8)

The Investigator (or delegate) RO/RN will conduct the following procedures:

- Adverse event review
- Concomitant medication review

#### Monthly research assessment (W4 and W8)

- Smoking status
- Quit attempts / Relapse
- Heaviness of Smoking Index (if still smoking)
- Substance use & wellbeing (ATOP)

The Investigator (or delegate) will provide the study medication as follows:

Condition 1: VNP

− Eight (8) bottles of 12mg/ml e-liquid
Condition 2: NRT (provided as below or equivalent)

− 28 x patches 21mg
− 2 x 20 pack of Inhalator 15mg
− 4 x Lozenge 4mg
− 1 x Gum (large) 4mg
A window of -7 days / + 14 days will be allowed for these visits.

#### End of Intervention (Week 12)

The RO/RN will conduct the following procedures:

- Adverse events review
- Concomitant medication review
- Advise/remind participant about upcoming CATI team call

An independent person from the CATI team shall conduct the following research interview over the phone.

In the event that the CATI Team has exhausted the maximum number of attempts to contact a participant, the site research team member may contact the participant and complete the interview.

#### End of Treatment research assessment

- Smoking status
- Quit attempts / Relapse
- Self-report 7-day point prevalence abstinence
- CO breath test if self-report 7-day abstinence
- Heaviness of Smoking Index (if still smoking)
- Craving, frequency and strength
- Minnesota Withdrawal Scale
- Quit intentions / motivation / difficulty / self-efficacy
- Amount spent on tobacco in the last week (if smoking)
- Attitudes to VPN (Condition 2 only)
- Substance Use & wellbeing (ATOP)

A window of -7 days / + 14 days will be allowed for this interview.

All participants who complete the End of Treatment interview will receive a $50 gift card.

An additional $50 gift card will be provided to participants who report 7-day abstinence from tobacco at this interview and complete a CO breath test for verification purposes.

#### Carbon Monoxide Testing

For all participants who self-report abstinence over of the past 7 days, by responding “No” to the self-report abstinence question: “Have you smoked at all, even a puff, in the last seven days?” the Investigator (or delegate) will, where possible, obtain a breath CO analysis within 14 days of being notified that a participant is eligible.

This may be done in person in the clinic where breath CO samples are permitted. Participants will be asked to attend their clinic where the research staff member will supervise the CO test. The Jarvis protocol will be followed where participants are asked to take a breath in, hold for 15 seconds and exhale into a breathalyser (Bedfont Smokerlyzer®) through a mouthpiece. The breathalyser provides an on-screen reading which is used to determine smoking status. In circumstances where clinic visits are not permitted, CO verification may be completed remotely by the participant using a personal CO monitor (e.g. iCO™ Smokerlyzer®) provided by the research team.

#### End of Treatment

A participant will be defined as having completed treatment once they have completed their end of Intervention (12-week) assessment, or at the time that they are prematurely withdrawn from treatment.

For all participants, including those that prematurely withdraw from treatment, all attempts must be made to carry out the End of Intervention interview as this is the measure for the primary endpoint of the study.

#### Follow up call reminder

As there is no formal research contact between week 12 and 24 interviews, site research staff are able to call a participant prior to their 24-week follow up to remind them of the upcoming CATI call.

#### Continuation of nicotine products after treatment

For participants in either group who have been unable to stop smoking tobacco in the intervention period and wish to continue treatment using e-cigarettes, a script for nicotine e-liquid may be supplied by the Investigator (or delegate) who must be an approved prescriber of liquid nicotine receive get written informed consent to do so as per current TGA guidelines.

NRT may also be provided to participants from either group after the 12-week intervention period to assist cessation and relapse management.

Both treatment options are at the discretion of the Investigator (or delegate), who is responsible for ensuring that continued treatments are documented accordingly in source notes.

### Follow Up

A follow up interview will be conducted at 3-months post treatment over the phone by a member of the independent CATI team. In the event that the CATI Team has exhausted the maximum number of attempts to contact a participant, the site research team member may contact the participant and complete the interview.

#### Follow up interview (Week 24)

- Smoking status
- Quit attempts / Relapse
- Self-reported 7-day point prevalence abstinence from tobacco smoking
- Self-reported continuous abstinence since last follow up (Week 12)
- Heaviness of Smoking Index (if still smoking)
- Craving, Frequency and Strength
- Minnesota Withdrawal Scale
- Quitting self-efficacy / motivation / difficulty / self-efficacy
- Current Nicotine product use
- Amount spent on tobacco in the last week
- Substance Use & wellbeing (ATOP)
- Current OAT treatment (medication, route of administration, dose, length of current treatment episode)

Participants that are unable to be contacted for the follow up interview within at least 45 days past the due date will be defined as lost-to-follow-up.

All participants who complete the follow-up interview will receive a $50 gift card.

#### End of Study

A participant will be defined as having completed the study once they have completed their 3-month post treatment follow-up assessment (Week 24-28); or if they are prematurely withdrawn from the entire study (i.e. treatment and research visits).

### Treatment discontinuation

Treatment discontinuation does not equate withdrawal from the study and these participants will be invited to continue completing the research component of the study.

#### Involuntary discontinuation

Participants may be withdrawn involuntarily by the investigator (or delegate) if they meet the following criteria:

- Participant experiences a severe or serious adverse event, thought to be related to the study drug/device, which is not resolving.
- Absence from the protocol monthly visits (Week 4, 8 or 12) for more than four weeks.

Even if the participant has capacity to consent at baseline, the intervention staff will assess capacity to consent at each contact and will cease the intervention if the participant is deemed to lack capacity. In this case the reasoning will be explained and recorded on the CRF.

#### Voluntary discontinuation

Participants have the option to stop treatment or revoke their consent at any time without giving a reason. The distinction between stopping treatment and revoking consent is shown by the following definitions:

- Discontinuation of treatment: A participant would be considered to have discontinued protocol treatment if they stop using the VNP device/NRT medication study. In this case the participant will continue in all remaining research interviews and assessments.
- Revocation of consent: Total withdrawal from the trial would occur in the circumstance that the participant decides to revoke their consent. Under these circumstances no further information would be collected from the participant for the purpose of the trial.

Participants may at any time elect to revoke consent for study participation without jeopardising their relationship with either their doctor or treatment service.

## STUDY ASSESSMENTS AND QUESTIONNAIRES

Assessments may be completed F2F, via telehealth/phone, or may be emailed to participants as surveys to be completed by them.

| **SCREENING ASSESSMENTS** |  |
| --- | --- |
| Medical history | At the screening visit a study doctor will record a medical and psychiatric history based on patient interview and physical examination, with emphasis upon active conditions and significant conditions within past 5 years |
| Substance use history | A survey of historical and current use of substances, including past month use, amount used per day previous treatment (yes/no), taken by study doctor. |
| Clinical assessment, vital signs | Sitting systolic and diastolic blood pressure, heart rate and weight will be obtained at Screening. |
| **BASELINE & OTHER ASSESSMENTS** |  |
| Demographics | Age (birthdate), gender identity, Aboriginal status, living arrangements, education and income will be recorded |
| Contact information | Participant's personal contact details and those of secondary contacts (e.g. family members, partners, and friends) shall be collected at the screening visit. |
| Concomitant medication | Concomitant medications shall be assessed and recorded from signing the consent form until Week 12. This includes all nicotine replacement treatments and devices administered (prescribed or self-administered). Any changes in concomitant therapy during the study will also be documented, including cessation of therapy, initiation of therapy and dose changes. |
| Current OAT treatment | Opiate treatment, including medication, dose, type and length of current treatment episode will be collected at Baseline and Week 24 to describe participant characteristics and outcomes. |
| Smoking and quitting history (including NRT use) | Smoking and quitting history including previous NRT use will be obtained at baseline as part of the researcher baseline interview questionnaire to examine effectiveness/predictability of smoking cessation success. |
| Quitting intention/ motivation/ difficulty/ self-efficacy | Predictors of successful quit attempt or relapse measured at baseline and Week 12 and 24. |
| Amount spent on tobacco in past week | To ascertain economic impact of smoking at baseline and Week 12. |
| VNP familiarity, use | Measured at Baseline and Week 12 to determine consequence of VNP use. |
| Substance use, health & wellbeing | The Australian Treatment Outcome Profile (ATOP) collects self-reported substance use, health and well-being data from the |
|  | preceding four-week period, for both clinical and research use and will be completed at screening, Week 4, 8, 12 and 24 |
| Patient Health Questionnaire-4 | Patient Health Questionnaire-4 is a 4 item screening tool for anxiety and depression in primary care settings. This will be assessed at baseline only for reporting of participant characteristics (30). |
| Co-use cannabis & tobacco | For those that use cannabis, amount of tobacco that is mixed, frequency and duration is assessed at Baseline for cannabis/tobacco co-use sub-study (see appendix 1) |
| Treatment preference | Participants will be asked which treatment (VNP or NRT) they would prefer if given a choice and how strong that preference is (Strong vs Moderate vs Weak). |
| **PRIMARY OUTCOME ASSESSMENTS** |  |
| Self-report 7-day point prevalence abstinence | 7-day point prevalence abstinence from tobacco smoking will be self-reported and based on the following dichotomous question pilot tested in the QuitNic trial (31), “Have you smoked at all, even a puff, in the last 7 days?” Completed at Week 12 |
| **SECONDARY OUTCOME ASSESSMENTS** |  |
| Carbon monoxide (CO) in expired air | Self-report 7-day point prevalence abstinence at end of treatment will be verified using a measure of carbon monoxide (CO) in expired air. Completed at Week 12. |
| Adverse Events | Adverse events shall be assessed and recorded from signing the consent form until Week 12. |
| Heaviness of Smoking Index (HSI) | The HSI measures time to the first cigarette of the day (5, 6–30, 31–60 and 61 + min) and average daily consumption of cigarettes (1–10, 11–20, 21–30, 31 + cigarettes (32). Completed at Screening and Baseline, Weeks 4, 8, 12 and 24. |
| Craving strength and frequency | Cravings to smoke will be assessed by one item based on Taggar et al (33) "Currently, how often do you get strong cravings to smoke tobacco?" with the response options of 1) Hourly or more often; 2) Several times per day; 3) At least once a day; and 4) Less than daily. Strength will be assessed by asking "How strong have these urges been?" with six response options ranging from No urge to Extremely Strong. Completed at Baseline, Week 12 and 24. |
| Nicotine Withdrawal - Minnesota Nicotine Withdrawal Scale | Nicotine withdrawal will be assessed by the Minnesota Nicotine Withdrawal Scale (34), an eight item scale rating withdrawal symptoms on an ordinal scale ranging from 0 (not present) to 3 (severe). Completed at Baseline, Week 12 and 24. |
| Quit attempts & relapse | Time to relapse and number of quit attempts will be assessed among those who relapsed. Completed at Baseline, Week 4, 8, 12 & 24. |
| Nicotine product adherence | Current use of short and long-acting NRT and e-cigarettes is assessed at Week 4, 8, 12 & 24. At weeks 12 & 24, additional questions around plans for long-term use, length of use and reasons for stopping use are also asked. |
| Self-reported 30-day continuous abstinence from tobacco smoking | 30-day continuous abstinence from tobacco smoking will be self-reported and based on the following dichotomous question pilot tested in the QuitNic trial (31) "Have you smoked at all, even a puff since the week 12 research interview?" The quantity of cigarettes smoked in the last 30 days will be assessed amongst those who smoked. Completed Week 24. |
| Self-reported 7-day point prevalence | Self-reported 7-day point prevalence abstinence from nicotine all (including NRT and VNP) will be measured by asking participants |
| abstinence from all tobacco smoking | "Have you smoked at all, even a puff, in the last 7 days?" Completed Week 24. |
| Health Economics<br>Cost Analyses | A cost consequence study setting out a detailed comparative cost of treatments - from the perspective of health care provider and; primary and secondary outcomes of the VNP and NRT. Costing items include public sources, wage awards, the Medical Benefits Schedule (MBS) and the Pharmaceutical Benefits Scheme (PBS); and research records such as costs of VNP products, inhalators, nicotine patches and gum, batteries and quitting aid materials.<br>Assessed at Week 12. |

8.2 Schedule of Assessments: Clinic & Research Visits

|  | Pre-screen | Screen | Base-line | Treatment |  |  |  |  |  |  |  |  |  |  | Follow up |
| --- | --- | --- | --- | --- | --- | --- | --- | --- | --- | --- | --- | --- | --- | --- | --- |
| COMPLETED BY | SITE Research staff |  |  |  |  |  |  |  |  |  |  |  |  | CATI | CATI |
| STUDY WEEK |  |  | W1 | W2 | W3 | W4 | W5 | W6 | W7 | W8 | W9 | W10 | W11 | W12 | W24 |
| WINDOW (days) |  |  |  |  |  | -7/+14 |  |  |  | - 7/+14 |  |  |  | -7/+14 | -7/+28 |
| ENROLMENT |  |  |  |  |  |  |  |  |  |  |  |  |  |  |  |
| Pre-Screening | X |  |  |  |  |  |  |  |  |  |  |  |  |  |  |
| Informed Consent |  | X |  |  |  |  |  |  |  |  |  |  |  |  |  |
| Eligibly Screening |  | X |  |  |  |  |  |  |  |  |  |  |  |  |  |
| Medical History |  | X |  |  |  |  |  |  |  |  |  |  |  |  |  |
| Randomisation |  | X |  |  |  |  |  |  |  |  |  |  |  |  |  |
| ALLOCATION |  |  |  |  |  |  |  |  |  |  |  |  |  |  |  |
| Provision of training |  |  | X |  |  |  |  |  |  |  |  |  |  |  |  |
| Set target quit date – set within 2 weeks |  |  | X |  |  |  |  |  |  |  |  |  |  |  |  |
| CONDITION 1 VNP: |  |  |  |  |  |  |  |  |  |  |  |  |  |  |  |
| • Provision of VNP/eLiquid (4-week supply) |  |  | X |  |  |  |  |  |  |  |  |  |  |  |  |
| • Provision of NRT (1-week supply) |  |  | X |  |  |  |  |  |  |  |  |  |  |  |  |
| CONDITION 2 NRT: |  |  |  |  |  |  |  |  |  |  |  |  |  |  |  |
| • Provision of NRT (4-week supply) |  |  | X |  |  | X |  |  |  | X |  |  |  |  |  |
| ASSESSMENTS |  |  |  |  |  |  |  |  |  |  |  |  |  |  |  |
| Demographics |  | X |  |  |  |  |  |  |  |  |  |  |  |  |  |
| Smoking status |  | X | X |  |  | X |  |  |  | X |  |  |  | X | X |
| Heaviness of Smoking Index (HSI) |  | X | X |  |  | X |  |  |  | X |  |  |  | X | X |
| Quit history |  |  | X |  |  |  |  |  |  |  |  |  |  |  |  |
| Quit attempts + relapse |  |  |  |  |  | X |  |  |  | X |  |  |  | X | X |
| Quit intention/motivation/difficulty/self-efficacy |  |  | X |  |  |  |  |  |  |  |  |  |  | X | X |
| Nicotine withdrawal (MTWS) |  |  | X |  |  |  |  |  |  |  |  |  |  | X | X |
| Craving frequency & strength |  |  | X |  |  |  |  |  |  |  |  |  |  | X | X |
| Amount spent on tobacco |  |  | X |  |  |  |  |  |  |  |  |  |  | X |  |
| VNP familiarity and use |  |  | X |  |  |  |  |  |  |  |  |  |  |  |  |
| Nicotine product adherence |  |  |  |  |  | X |  |  |  | X |  |  |  | X | X |
| Co-use cannabis & tobacco |  |  | X |  |  |  |  |  |  |  |  |  |  |  |  |
| Current opiate treatment |  |  | X |  |  |  |  |  |  |  |  |  |  |  | X |
| Substance Use (ATOP) |  | X |  |  |  | X |  |  |  | X |  |  |  | X | X |
| Health & wellbeing (ATOP) |  | X |  |  |  | X |  |  |  | X |  |  |  | X | X |
| Baseline health (PHQ-4) |  |  | X |  |  |  |  |  |  |  |  |  |  |  |  |
| Treatment Preference |  |  | X |  |  |  |  |  |  |  |  |  |  |  |  |
| **ABSTINENCE MEASURES** |  |  |  |  |  |  |  |  |  |  |  |  |  |  |  |
| 7-day point prevalence abstinence from tobacco |  |  |  |  |  |  |  |  |  |  |  |  |  | X | X |
| Breath CO test (within 14-days of eligibility notification) |  |  |  |  |  |  |  |  |  |  |  |  |  | X |  |
| 30-day continuous abstinence from tobacco |  |  |  |  |  |  |  |  |  |  |  |  |  |  | X |
| **CLINICAL ASSESSMENT, VITALS & SAFETY** |  |  |  |  |  |  |  |  |  |  |  |  |  |  |  |
| Vital Signs |  | X |  |  |  |  |  |  |  |  |  |  |  |  |  |
| Weight |  | X |  |  |  |  |  |  |  |  |  |  |  |  |  |
| Adverse Events |  |  | X |  |  | X |  |  |  | X |  |  |  | X |  |
| Concomitant Medications |  |  | X |  |  | X |  |  |  | X |  |  |  | X |  |
| **HEALTH ECONOMICS COST ANALYSIS** |  |  |  |  |  |  |  |  |  |  |  |  |  | X |  |

## RISKS

### Overview

VNPs with nicotine e-liquid and NRT are currently used products and services designed to help people quit smoking, however they have not been formally linked together in the population described for this study.

Risk of physical harm to the research assistant or service staff members during enrolment in the trial or completion of the assessments is unlikely.

The risk of physical harm during the intervention if the participant uses supplied NRT and/or VNP with nicotine e-liquid include the known side effects specified in section 9.1 and 9.2. Quitting smoking can also be associated with withdrawal symptoms and increased side effects of medications.

VNPs with nicotine e-liquid have been available for decades, however long-term safety data has not reliably indicated a significant safety risk. There is some indication of risk to physical harm associated with VNP with nicotine e-liquid use, such as cardiovascular problems. VNPs and nicotine e-liquid are considered to be less harmful than regular tobacco cigarettes.

There is also potential risk of participants using other liquids/THC or cannabis oil, including ‘street’ liquids in their VNP device. Use of such liquids in VNP devices has been associated with hospitalisations and death in the USA.

Doses of nicotine that are tolerated by adult smokers can produce severe symptoms of poisoning in small children and may prove fatal. E-liquid must be kept out of reach of children at all times and participants will be advised of this in product labelling and instructions for use.

Risk of psychological harm during quitting is unlikely. However, in planning for or quitting smoking, some participants may experience associated distress and/or discomfort. Assessment elements are standard and include self-report measures and carbon monoxide (CO) measurement. Standard assessment risks include risk of distress or discomfort from answering personal and sensitive questions. Some of the measures address symptoms of emotional distress and/or psychiatric symptomatology and there is a possibility that participants may experience some emotional discomfort whilst relaying this information.

There is also the risk of a breach of confidentiality (e.g. if information is required by law, or in the case of risk of harm to self or others).

The risk of inconvenience to participants will be minimal for those completing telephone follow-ups. For those participants having CO measurement at Week 12 or Week 24 there may be inconvenience involved in attending those interviews.

### Vaporised Nicotine Products

The combination of the Innokin Indura VNP with Nicophar or Lion Labs e-liquid has not formally been linked together before in the clinical trial setting or with the population described for this trial therefore a safety profile has not been established.

A Systematic Review of literature performed by McRobbie et al in 2014 (25) has identified the possible side effects in electronic cigarettes, with the most common being:

- Cough
- Dry mouth
- Throat irritation
- Mouth Irritation
- Headache
- Phlegm.

Other less common side effects were indigestion, gas, and diarrhoea, shortness of breath, irregular heartbeat, hiccups, dizziness, taste disturbance, dysgeusia (loss of taste), abdominal discomfort, nausea and vomiting.

### Nicotine Replacement Therapy

The most common side effects associated with Nicotine Replacement Therapy are;

- Vivid dreams
- Itchiness
- Swollen, red patch sites
- Dry mouth
- Dry throat
- Cough
- Nausea
- Headache.

## ADVERSE EVENT DEFINITIONS, RECORDING AND REPORTING

### Adverse Event

An adverse event (AE) is the appearance or worsening of any undesirable sign, symptom, or medical condition that does not necessarily have a causal relationship with the treatment.

Nicotine withdrawal symptoms will be captured on a separate assessment form (Minnesota Nicotine Withdrawal Scale) and should not be recorded as AEs.

Common Nicotine withdrawal symptoms include:

- Angry, irritable or frustrated
- Anxious or nervous
- Depressed or sad
- Desire or craving to smoke
- Difficulty concentrating
- Increased appetite, hunger or weight gain
- Insomnia, sleep problems or awakening at night
- Restlessness or impatience

The Investigator/Study Doctor will determine if a participant is experiencing Nicotine withdrawal symptoms if the above symptoms are reported.

The appearance of any other condition must be documented on the AE Case Report Form.

Information collected concerning AEs will include; name of event, onset date, resolution date, severity, expectedness, relatedness, seriousness, action taken (if any) and outcome of event.

#### Severity

The severity of an adverse event refers to the intensity of the event without regard to whether or not it meets the criteria for serious. Severity is expressed in grades:

Mild: ￼ Discomfort noticed but no disruption of normal daily activity

Moderate: ￼ Discomfort sufficient to reduce or affect normal daily activity

Severe: ￼ Incapacitating with inability to work or perform normal daily activity

Life threatening: ￼Represents an immediate threat to life

#### Expectedness

Expectedness is assessed using the trials reference safety information; the information contained in either an investigator’s brochure or an approved Australian Product Information (or another country’s equivalent) that contains the information used to determine what adverse reactions are to be considered expected adverse reactions and, on the frequency and nature of those adverse reactions. Expectedness is currently limited in its evidence for the use of e-liquid and e-cigarettes. An updated Cochrane review found no reported serious adverse events related to EC use, most frequently reported adverse events were mouth and throat irritation, usually decreasing over time. (25)

#### Relatedness

The reporting investigator must provide their opinion on the relationship of the AE to the allocated intervention (i.e. NRT or VNP). Relationship may be defined as:

Unlikely: ￼An adverse event that is unlikely to be related to the use of the allocated intervention

Possibly: ￼An adverse event that might be related to the use of the allocated intervention

Probably: An adverse event that is likely to be related to the use of the allocated intervention

AEs judged as possibly or probably caused by the allocated intervention would qualify as adverse reactions.

For queries or advice regarding determining the relationship of the AE to the VNP please contact Conjoint Associate Professor Colin Mendelsohn by contacting Project Manager for details.

#### Seriousness

Adverse events are considered ‘serious’ if they threaten life or function. Due to the significant information, they provide, Serious Adverse Events (SAEs) require expedited reporting.

SAEs are defined as any adverse event which:

- Results in death
- Is life-threatening (an event in which the participant was immediately at risk of death at the time of event)
- Results in a persistent or significant disability/incapacity
- Results in a congenital anomaly/birth defect
- Requires inpatient hospitalisation or prolongation of existing hospitalisation
- Results in a medically important event or reaction.

Medical and scientific judgment should be exercised in deciding whether other situations should be considered serious, e.g. important medical events that may not be immediately life threatening or result in death or hospitalisation but may jeopardise the participant or may require intervention to prevent one of the outcomes listed in the definition above. This definition also includes serious adverse clinical consequences associated with use outside the terms of the protocol including, for example adverse device effects arising from the use of cannabis oil in the VPN.

An event will not be considered to be an SAE if:

- Hospitalisation is for routine treatment or monitoring of the studied indication, not associated with any deterioration in condition
- Hospitalisation is for elective or pre-planned treatment for a pre-existing condition that is unrelated to the indication under study and has not worsened since the start of study drug
- Treatment is provided on an emergency outpatient basis for an event not fulfilling any of the definitions of an SAE given above and not resulting in hospital admission
- Hospitalisation is for social reasons and respite care in the absence of any deterioration in the patient’s general condition is medically significant, i.e. defined as an event that jeopardizes the patient or may require medical or surgical intervention to prevent one of the outcomes listed above.

### Adverse Device Effects

Is an adverse event related to the use of an investigational medical device?

Note: This definition includes adverse events resulting from insufficient or inadequate Instructions for use, deployment, implantation, installation, or operation, or any malfunction of the investigational medical device. This definition includes any event resulting from use error or from intentional misuse of the investigational medical device.

The intentional misuse of cannabis oil or any other non-prescribed liquid in the vaporised nicotine product is reportable as an Adverse Device Effect.

The site investigator will determine if an Adverse Device Effect has occurred, and the expectedness based on the reference safety information.

### Device Deficiency

Inadequacy of a medical device with respect to its identity, quality, durability, reliability, safety or performance.

Note: Device deficiencies include malfunctions, use errors, and inadequate labelling.

The site investigator will determine if a device deficiency event has occurred based on information received from the participant and the reference safety information.

### Serious Adverse Device Effect (SADE)

An adverse device effect that has resulted in any of the consequences characteristic of a serious adverse event. Any SADE is required to be reported to the Project Manager within 24 hours of becoming aware of the event.

### Unanticipated Serious adverse device effect (USADE)

A Serious Adverse Device Effect which by its nature, incidence, severity or outcome has not been identified in the current version of the risk analysis report. Any USADE will have the same reporting requirements and deadlines specified in section follow the same reporting timeframes as a SUSAR covered in section 10.2.2 below.

Note: Anticipated serious adverse device effect (ASADE) is an effect which by its nature, incidence, severity or outcome has been identified in the risk analysis report.

### Adverse Event Reporting

#### Adverse Event/Device/Reaction Reporting

AEs are reported from the time the participant has signed the consent form until at least 30 days after the end of treatment (Week 12 or early discontinuation) must be recorded on the AE Case Report Form.

AEs though to be related to the investigational product/s will be classified as Adverse Reactions (ARs). This trial will adhere to the NHMRC safety and reporting in clinical trial guidance document (35)

#### Serious Adverse Event/Device Reporting

AEs/ADEs classified as serious (SAEs) during treatment up until 30 days after end of treatment must be report to the Project Manager WITHIN 24 HOURS OF KNOWLEDGE OF THE EVENT by telephone or email using the Serious Adverse Event reporting form.

When all information is not included in the initial report, a detailed follow up report must be provided WITHIN THE NEXT 24 HOURS.

The immediate and follow up reports should identify participants by the participants unique study ID rather than personal identification. The Investigator must also comply with applicable institutional governance requirement/s relating to the reporting of serious adverse events.

Any serious adverse event that is ongoing at the end of study (i.e. 3-month follow-up) must be followed until resolution or until the event stabilizes (for those events that will not resolve).

SAE reports must be submitted to the Project Manager according to the instructions provided on the SAE report form.

#### Suspected Unexpected Serious Adverse Reaction Reporting

Adverse Events that that are suspected to be UNEXPECTED according to the reference safety information, RELATED to the allocated intervention and SERIOUS are classed as Suspected Unexpected Serious Adverse Reactions (SUSARs).

The SAE reporting requirements described above must be followed by Investigators reporting SUSARs (i.e. report to the Project Manager WITHIN 24 HOURS OF KNOWLEDGE OF THE EVENT BY TELEPHONE OR EMAIL USING THE SERIOUS ADVERSE EVENT REPORTING FORM).

The Project Manager (by delegation of the Trial Chairpersons) will coordinate expedited reporting of any SUSAR to the IDSMC, all concerned investigators/institutions, Therapeutic Goods Association, HRECs, and regulatory authorities within the reporting timeframe.

Reports will comply with applicable regulatory requirements and ICH Guideline for Clinical Safety Data Management: Definitions and Standards for Expedited Reporting.

## PREGNANCY

Participants who become pregnant will be withdrawn from the study (as identified in the exclusion criteria). NRT will be provided as a duty of care, nicotine delivered via these products is safer than continued smoking during smoking pregnancy. (36) Follow-up information will be obtained where possible regarding the course and outcome of the pregnancy.

## PACKAGING, LABELLING, STORAGE, AND ACCOUNTABILITY OF NICOTINE PRODUCTS

### Formulations

#### Innokin Endura T18-II

Innokin Endura T18-II, a refillable tank style device, will be used for this study to deliver liquid nicotine. This simple to use, reusable device has the ability to deliver nicotine more efficiently than disposable devices.

The Innokin Endura T18-II device was selected based on manufacturer compliance with Good Manufacturing Practice standards.

#### Liquid nicotine

Nicophar liquid nicotine (or e-liquid) 12mg/mL, in 10ml bottles, will be used with the Innokin Endura T18-II. Each 10mL bottle will contain nicotine (1.2%), Glycerol (84%) and water (14.8%).

Nicophar liquid nicotine was selected based on manufacturer compliance with Good Manufacturing Practice standards.

Lion Labs liquid nicotine will be made available to replace Nicophar liquid. Manufactured in New Zealand to meet current TGA approved safety and packaging requirements, the 12mg/mL 10ml bottles each contain nicotine (1.2%), vegetable glycerine (82.8%) and purified water (16%)

#### Nicotine Replacement Therapy

The following forms of Nicotine Replacement Therapy will be used for this study:

- Patches 21mg
- Inhalators 15mg
- Lozenge 4mg
- Gum 4mg
- Oral mouth spray 1mg/spray.

### Packaging and Labelling

#### Innokin Endura T18-II

The Innokin Endura T18-II device is provided as part of a starter kit also containing 1 x Endura T18 II Battery (1300mAh), 1 x Prism T18 II Tank, 2 x 1.5Ω Coils, 1 x Magnetic Cap, 1 x Spare Drip Tip, Spare O-Rings, 1 x USB Charging Cable and a Quick Start Guide. An additional 5 x 1.5Ω Coils will be supplied separately making 7 coils in total.

#### Liquid nicotine

Labelling of nicotine will be conducted by the participating sites pharmacy and should comply with Drugs, Poisons and Controlled Substances Regulations 2006, the SUSMP, and Annex 13 of the Clinical Trial Handbook. Doses of nicotine that are tolerated by adult smokers can produce severe symptoms of poisoning in small children and may prove fatal.

E-liquid must be kept out of reach of children at all times and participants will be advised of this in product labelling and instructions for use. Child resistant closures will be supplied on the bottles.

#### Nicotine Replacement Therapy

NRT kits will be made up by HARMONY research members at the University of Newcastle and include the product’s original packaging. As stated above, precautions need to be made around the correct storage and disposal of NRT around children. This will be stated in the instructional guidance included in the packaging, and participants will be advised when dispensed.

### Deficient or lost devices

Should devices be found to be deficient, a replacement will be sent to the participant. The deficient device must be returned to either the site or coordinating centre for return to manufacturer. Should a device be stolen, lost or damaged by participants a replacement may be offered on the first instance only and will be at the discretion of the site investigator.

### Dispensing and Accountability

**Condition 1:** VNP intervention packs (containing study device, device information materials, 1-week supply of NRT patches) will be stored securely at the University of Newcastle located within the McAuley Building, Calvary-Mater Hospital, Waratah. These will be supplied to each of the participating services pharmacy for appropriate labelling, storage, and dispensing as per site pharmacy procedures. In addition to pharmacy records, research staff will maintain a log of study products dispensed to participants including recording batch numbers in CRFs.

**Condition 2:** NRT will be packaged into kits and stored securely as above, prior to being sent to participating sites. No scripts or additional labelling will be required for dispensing to participants and NRT dispensed will be recorded by research staff in CRFs.

### Study drug destruction

Condition 1: All VPNs and e-liquid dispensed to the participant will be kept by the participant at the end of the study. There will be no provisions for the return of the VPN and e-liquid. Any remaining e-liquid will be destroyed as per local pharmacies existing operating procedures.

Condition 2: NRT dispensed to participants will be kept by the participant. There will be no provisions for the return of NRT. Any remaining/expired NRT will be returned to the lead site for distribution as directed by study investigators or destroyed as per local pharmacies existing operating procedures.

## DATA QUALITY ASSURANCE

### Regulatory Documents and Records Retention

The Principal Investigator is responsible for creating and/or maintaining all study documentation required by ICH GCP section 8, as well as any other documentation defined in the protocol. The Investigator must provide key documents to the Project Manager/Trial Coordinating Centre prior to the start of the study. A complete list of required regulatory documents will be supplied by the Project Manager/Trial Coordinating Centre. ICH GCP Guidelines require that the Investigator retain a copy of all regulatory documents and records that support the data for this study for 15 years after completion of the trial.

If a Principal Investigator should retire, relocate or for other reasons withdraw from the responsibility of keeping the study records, custody will be transferred to the replacing Principal Investigator.

### Adherence to Protocol and Good Clinical Practice

This study will be conducted under Good Clinical Practice (GCP) and as per the terms and instructions described in the approved protocol. Any breach, divergence or departure from the requirements of the protocol or GCP must be reported as a ‘deviation’. Each deviation will be assessed by the Trial Chairperson (or delegate) for seriousness.

Exceptions may be made for:

a. An emergency situation in which proper care for the protection, safety and wellbeing of the trial participant requires that an alternative treatment be used, or
b. A planned, one time, temporary action requested via a Protocol Exception Form.

#### Deviations

All deviations shall be assessed for seriousness. The term serious breach is defined by the NHMRC Reporting of Serious Breaches of Good Clinical Practice (GCP) or the Protocol for Trials Involving Therapeutic Goods, 2018 as an in incident or occurrence that is likely to affect to a significant degree:

a. The safety or rights of a trial participant, or
b. The reliability and robustness of the data generated in the clinical trial.

The Trial Chairperson or their delegate will report serious breaches directly to the reviewing HREC as per the NHMRC Reporting guidelines.

#### Exceptions

A protocol exception is a planned, one-time, temporary action or process that departs from the ethics approved protocol, and generally intended for one specific participant.

An exemption request will only be considered if it does not impact the rights, welfare or safety of present, past or future participant(s); increase the risks and/or decrease the benefit for research participants, compromise the integrity of the study data, or affect the participants willingness to participate in the study.

Protocol exceptions shall be sought from the Chief Investigator (or delegate) by completion and submission of the study specific Protocol Exception Form.

### Data Collection and Security

This study will use electronic data capture in the form of REDCap (Research Electronic Data Capture). The REDCap database is located on a standalone database server hosted by Hunter New England Local Health District (HNELHD). The database server resides behind the HNELHD internal firewall and access to the server is controlled via firewall rules. All data collected via REDCap is backed up daily, both on the local server and by the HNELHD backup system. All connections to the system, both external and internal, will occur over encrypted channels.

Access to study records within the study database has been limited by using Data Access Groups (DAGS). Only users within a given DAGs can access records created by users within that group. Access to components of study records is role-based and can only be granted by the Project Manager of the study.

All data entered into REDCap for the purpose of data analyses will be de-identified and traceable to supporting identifiable source documentation such as hospital/medical records (including electronic health records), laboratory results, data recorded in automated instruments and pharmacy records, etc. All source documentation will be held confidentially in line with current legislation governing health information, and will not be made publicly available.

All study documentation (electronic and paper copies) will be archived securely at the end of the study for 15 years and then destroyed by secure destruction

### Data Sharing Plan

As submitted with the Australia New Zealand Clinical Trials Registry, data resulting from HARMONY will be shared in accordance with the following parameters:

- All of the data collected will be de-identified
- Data will be available beginning 3 months and ending five years after the Protocol is published
- Data will be available to those providing a sound methodology
- The data will be approved to meet the aims of the approved study only
- Proposals should be directed to Prof Adrian Dunlop and to gain access, requestors will need to sign a data access agreement with Hunter New England Local Health District.

### Study Monitoring

Monitoring of the study site may be performed by the Project Manager or designee. A risk assessment has determined that this is Type A (low risk) trial therefore a centralised monitoring approach will be adopted. De-identified CRFs will be uploaded to the REDCap database for remote verification of primary endpoints, eligibility, protocol adherence and data quality. Site visits are not planned but may be triggered by concerns identified from centralised monitoring that cannot be addressed by other means.

By signing the protocol, the Investigator agrees that, within local regulatory restrictions and institutional and ethical considerations, authorized representatives may review the site source documentation remotely to verify against data recorded in the database.

### Data Entry Timelines

To ensure safety and/or data quality issues are identified or detected in a timely manner data must be entered into the REDCap study database during collection or as soon as possible after collection, preferably within 3 business days.

Data queries raised in REDCap and/or sent via email must be addressed within 5 business days.

## STATISTICAL CONSIDERATIONS

### Sample Size

Studies using continuous abstinence measures with mental illness and AOD samples have found smoking cessation rates close to zero at longer term follow-up using NRT. (37) Using the same outcome measures, Bullen & Walker (38) found no significant difference in continuous abstinence between VNP and NRT groups (7.3% vs 5.8% respectively at 6 months follow-up) in a general population sample in New Zealand. Thus, for this trial, with our sample of heavy smokers with AOD use, we will conservatively assume a verified continuous abstinence rate of 1% at 6 months follow up in the NRT control group. A sample of 200 smokers in each treatment group are needed to detect a difference of 6% between groups (i.e., 1% in NRT group and 7% in VNP group continuous abstinence at 6-month follow-up) with 80% power and a 5% significance level. Based on our previous research in AOD setting with clients from various programs including OAT and assuming 30% attrition rate at 6 months follow-up, we will require a sample size of 572 eligible smokers across 6 OAT sites.

### Outcome Measures and Analysis Plan

#### Primary Outcomes

1. To examine the effectiveness of treatment with VNP compared to best practice NRT on self-reported 7-day point prevalence abstinence (from tobacco smoking) at the end of 12 weeks of nicotine treatment in OAT clients.

#### Secondary Outcomes

2. Difference between the proportions of participants in the VNP vs NRT groups for biochemically verified continuous abstinence measured via a CO monitor breath test for participants who self-report 7 days of continuous abstinence at end of treatment (Week 12). CO ppm used to assess abstinence will be >8pm.
3. Difference in the number, severity, expectedness, relatedness and seriousness of Adverse Events (including Adverse Device Events) in the study population.
4. Difference in the proportion of participants in the two treatment groups (VNP vs NRT) who self-report continuous abstinence from tobacco smoking since the Week 12 research interview at 3 months post-treatment (Week 24).
5. Difference in the proportion of participants in the two Treatment Groups (VNP vs NRT) who self-report 7-day point prevalence abstinence from tobacco smoking at 3 months post-treatment (Week 24).
6. The difference in the number of tobacco cigarettes smoked per day at baseline vs end of treatment (Week 12) and 3 months post-treatment (Week 24) will be compared for participants in the two treatment groups (VNP and NRT).
7. The difference in the nicotine craving and withdrawal symptoms at end of treatment (Week 12) and 3 months post-treatment (Week 24) relative to baseline will be compared for participants in the two treatment groups (VNP and NRT).
8. Difference in the number and duration of relapse episodes between the two treatment groups (VNP vs NRT) at end of treatment (Week 12) and 3 months post-treatment (Week 24) .
9. Difference in VNP/NRT treatment adherence between the two treatment groups at end of treatment (Week 12) and 3 months post-treatment (Week 24).
10. Difference in other substance use between the two treatment groups at end of treatment at end of treatment (Week 12) and 3 months post-treatment (Week 24).
11. Difference in study retention between the two treatment groups (VNP vs NRT) at end of treatment (Week 12) and 3 months post-treatment (Week 24).
12. Comparison of the cost and effect between VNP and NRT, where effect is the trial’s primary outcome: the proportion of people self-reporting 7-day point prevalence smoking abstinence at the end of treatment (Week 12).

Complete details of the statistical analyses to be performed will be documented in a statistical analysis plan (SAP), which will be completed prior to database lock. This document will include more detail of analysis populations, summary strategies, and any amendments to the proposed analyses, if necessary. Any changes to the SAP will be outlined in the final trial report.

### Early Termination

An Independent Data Safety Monitoring Committee may recommend stopping the trial should the number and/or severity of adverse events justify discontinuation of the study.

In the case of early termination of the study, the Trial Chairperson will advise the HREC, study sites and appropriate regulatory authorities. Principal investigators at each site will be responsible for informing participants at that site.

## RESEARCH GOVERNANCE

### Trial Management Committee

The Trial Management Committee (TMC) will be responsible for monitoring of the progress of the trial, decision-making, education and information services and reporting as described in the HARMONY *Trial Management Committee Terms of Reference*. TMC members are listed at the beginning of the protocol.

### Independent Data Safety Monitoring Committee

An Independent Data Safety Monitoring Committee (IDSMC), separate to the TMC, will independently monitor the conduct of the trial in order to ensure its ethical and scientific integrity. The roles and responsibilities of the IDSMC are set out in the *IDSMC Terms of Reference* document.

## PARTICIPANT PROTECTION

### Privacy and Confidentiality

The Principal Investigator is responsible for complying with applicable privacy regulations, per his or her state. Only information identified in this protocol will be collected. The information collected will only be used for the purposes identified in this protocol.

To ensure anonymity and to limit disclosure, participants will be assigned a unique Study ID. An identifier log will be maintained, linking each participant name to the corresponding identifier. Participant Enrolment Logs can be generated through the REDCap database.

Instructions are included in the Manual of Operations. Client confidentiality has been ensured by limiting access of these logs to site specific DAGs.

The results of this study will be reported in such a manner that participants will not be identifiable in any way. Published reports or presentations will refer to grouped data or coded individual data and not to any identifiable individuals.

By signing the Consent Form, the participant consents to the collection, access, use, and disclosure of his or her information as described in the Participant Information Sheet.

By signing this protocol, the Investigator affirms that he or she will maintain in confidence information furnished to him or her by the Trial Chairperson and will divulge such information to his or her respective HREC under an appropriate understanding of confidentiality with such board.

### Insurance and Compensation

The Treasury Managed Fund (TMF), a self-insurance scheme of the New South Wales (NSW) Government, provides indemnity and/or insurance coverage in relation to clinical trial activities conducted at public institutions in NSW

In the unlikely event that a trial participant suffers injuries or complications as a result of this study they may have a right to take legal action to obtain compensation. If the participant is not eligible for compensation under the law, but are eligible for Medicare, they can receive any medical treatment required free of charge as a public patient in any Australian public hospital.

## PUBLICATION AND PRESENTATION POLICY

The Trial Chairperson and TMC are responsible for presentations and publications arising from this trial. Refer to the *HARMONY Authorship, Publication and Spokesperson Guidelines* for detailed information.

## Data Availability

All data produced in the present study are available upon reasonable request to the authors

## APPENDIX 1 - CANNABIS AND TOBACCO CO-USE SUB STUDY

### Consumer driven treatment options for co-occurring cannabis and tobacco use: collaboration with Hunter opiate treatment clients

### Background

Cannabis and tobacco use commonly co-occur (39). This is known as co-use and refers to either the use of both independently (either at the same time or on different occasions) and/or the use of both substances mixed together (40). In Australia, the use of both substances is particularly evident in people with drug and/or alcohol concerns (41). This is a group at high-risk of early death from smoking-related causes and who are often overlooked for tobacco treatment (42).

The complex relationship between the two substances also compromises attempts by consumers to stop using tobacco or cannabis alone (43). Cessation of one substance often leads to increased use of the other and vice versa (44). Furthermore, co-use makes verification of abstinence difficult in tobacco smoking cessation studies (45), so treatments that address both substances would be well placed in tobacco treatments for alcohol and other drug populations.

Evidence suggests that concurrent tobacco and substance use treatment improves long-term abstinence outcomes (46). Despite this, treatment options for co-occurring cannabis and tobacco use are scarce. Effective intervention development requires an in-depth exploration of consumer experiences co-using these substances, their willingness to quit one or both, perceived barriers to cessation and treatments that may assist quitting. This feedback will provide a roadmap for an innovative and client-centred approach to cessation.

#### Aims

The study aims to explore concurrent use of cannabis and tobacco amongst opiate treatment clients and examine simultaneous cessation of both substances. Specifically it will examine:

- The prevalence of co-use in NSW opiate treatment populations
- Consumers’ quit histories and preferences around cessation of both substances
- Desire to quit both substances simultaneously
- Explore interventional strategies that may assist them to stop using both substances together

The information generated will be used to develop the framework for a novel cannabis and tobacco cessation intervention.

### Method

#### Study Design

The study will use a mixed-methods research design. The quantitative portion of the study will use data collected during research interviews conducted with Harmony participants who report cannabis use. This data will be captured electronically on the Harmony study REDCap database.

The qualitative portion of the study will involve in-depth interviews with up to 30 participants. These will be undertaken by a member of the research team. Interviews will be recorded and should take approx. 30 minutes to complete.

#### Participants

The study will utilise participants of the Harmony study who are clients of opiate treatment services and who currently smoke cigarettes. Those who self-report cannabis use will be asked questions about their use of tobacco and cannabis together.

A subset of participants from the Hunter New England and Sydney Local Health District sites will be asked about their interest in participating in an in-depth interview. Those interested will be contacted by a member of the HNE Drug and Alcohol Research Unit, the coordinating site for the Harmony study, and undergo a separate consent process prior to interview commencement. It is anticipated that interviews will be conducted face-to-face, but in circumstances where this is not possible, interviews will use audio or video conference technology.

It is anticipated that up to 30 clients will complete interviews, however, recruitment will continue until saturation or redundancy or data collection is reached.

Participants will be reimbursed for their time and receive a $25 retail gift voucher.

#### Outcomes

In addition to demographic and substance use information already collected, the quantitative portion of the study will examine the following:

- Prevalence of co-use in opiate treatment populations
- Prevalence of mixing tobacco with cannabis in these groups
- Simultaneous use of tobacco and cannabis including quantity and frequency
- History of cannabis quit attempts
- Whether participants would like to stop smoking cannabis and cigarettes and if so, whether they would consider stopping them simultaneously
- Whether participants believe there is a need for treatment that targets smoking and cannabis use together
- Whether co-use changes with the introduction of vaping for smoking cessation

The qualitative interviews will ask open-ended questions to gain an understanding of:

- Participants experiences of tobacco and cannabis use, including exploring users likes and dislikes and perceived benefits such as stress relief and sleep
- Past attempts at quitting either substance
- Reasons for wanting to quit both substances
- Willingness to quit both substances at once
- Perceived barriers and enablers to cessation of both substances at once
- Ideas about strategies that could assist cessation of both substances at once.

### Confidentiality

For those taking part in in-depth interviews, permission will be sought to audio-record the interview to allow for transcription and analysis of the interview material. During this process, all identifying information will be removed. Quotes from the discussion may be used in written reports without any distinguishing details included.

Participants will be informed during the consent process about confidentiality and that all information provided by them will remain confidential within the bounds of mandatory reporting and the law.

### Analysis

Descriptive statistics will be employed to analyse quantitative data that will primarily consist of: demographic information (including age, gender, Aboriginal status, education, income and household members), smoking data, substance use, and questions relevant to this study.

The qualitative data will be transcribed as soon as possible after interviews and entered into the software program NVivo 10 to assist with analysis. A coding book will map interview questions and this will be used to examine the data using qualitative description, a qualitative methodology commonly used in health research that draws from a natural perspective and provides rich descriptions of the perceptions and experiences of informants.

Two researchers will complete the coding, using discussion to agree on coding and any emerging descriptive categories. Analysis will be completed using Iterative Categorisation (Jo Neale Ref), a systematic and transparent data management technique developed in the field of addiction research.

### Reporting and publication

Data confidentiality, integrity and security will be as per the main protocol.

Results of this study will be used in their own right and in conjunction will data collected from the main study to inform the feasibility of an intervention targeting co-use of tobacco and cannabis in opiate treatment clients.

The investigators plan to discuss/publish the study findings of this research component in publicly available reports, journals, at relevant conferences and other appropriate public forums. Information will be provided in such a way that the participant cannot be identified. No personal details will be released in any publication.

## APPENDIX 2 - PARTICIPANT EXPERIENCES OF VAPORISED NICOTINE PRODUCTS

### Background

The results of the Harmony clinical trial will provide comprehensive evidence for the effectiveness of vaporised nicotine as a tobacco treatment strategy relative to nicotine replacement therapy. It does not however, provide information on participants’ perspectives and experiences utilising vaporised nicotine as an aid to smoking cessation. Patient perspective information would complement the overall study results and provide a more holistic picture of vaporised nicotine products (VNPs) as a tobacco treatment option for opiate treatment clients.

### Aims/Hypothesis

This sub study aims to understand the patient experience of and attitudes towards VNPs as a means to achieve smoking cessation in opiate agonist treatment patients.

To do this, it will investigate the perceived benefits and challenges associated with VNPs as a tool for smoking cessation plus self-reported effectiveness and compliance with vaporised nicotine in participants enrolled in the main Harmony study.

### Methods

#### Study design

The sub study will utilise qualitative methods to achieve its aims. Data collection will involve in-depth interviews with study participants using pre-prepared question templates.

#### Participants

Participants will be those enrolled in the main study who were randomised to the VNP treatment group and have completed the 12-week treatment phase of the study.

#### Procedures

Eligible participants will be identified and contacted by Harmony research staff, who will explain the sub study aims and its procedures. If participants are interested in taking part, an appointment will be made with the option of phone or face to face interviews.

Participants will undergo a separate consent process with the researcher prior to interview commencement. It is anticipated that up to 30 participants will complete interviews, however, recruitment will continue until saturation or redundancy of data collection is reached.

It is expected that the interviews will take approximately 20 minutes to complete. Participants will be reimbursed for their time and receive a $20 retail gift voucher.

### Semi-structured Interviews

#### Before treatment

“We’d like to know about your experience smoking tobacco before you became involved in this study…”

1. “What did you smoke and how often?”
2. “Have you tried quitting before?

a. If yes, with what and how did it go?
b. If no, what made it difficult?”
3. “What do you think kept you smoking tobacco?”
4. “Had you tried vaping nicotine before you joined this study?

a. If no, why not?
b. If yes, why didn’t you continue vaping?”
5. “Before you joined the study, were you hoping to be in the Vape or NRT group?”
6. “What did you think the chances were that vaping would help you to stop smoking?”

#### During treatment

##### Implications of vaping for nicotine use

“Now we’d like to know about your experiences during the study, with vaping nicotine…”

7. “What did you like and dislike about vaping?”
8. “What was it like learning to use the device?”
9. “Were there things that made it easy or difficult to use?”

a. Did the device work ok?
b. Did you experience a pleasing nicotine effect?
10. “Did you experience any nicotine withdrawal or side effects?

If so, can you tell us more about these?”

11. “Compared with smoking, did vaping have any effect on your nicotine cravings?”

##### Implications for Opioid use

“Now we would like to know how vaping affected your opioid or other drug use”

12. “Once you joined the study and started vaping, did you find yourself using other methods to relieve cravings (e.g., using heroin/other drugs, relaxation to reduce stress)?

##### Interactions with Opiate Treatment

13. “Some patients find that their desire to smoke increases shortly after receiving methadone or bupe. Did you experience this with your smoking?

a. Did you find that you smoked more?
b. How long did that last?”
14. “Since you’ve been vaping, have you noticed similar effects?

a. If yes, how does it compare to the effect of smoking?”
15. “What is similar about being on buprenorphine or methadone (to cut back on heroin or other opioids) and vaping nicotine (to cut back on tobacco) for you?”

a. Do you worry you might be swapping one type of addiction (smoking tobacco) for another (vaping nicotine)?
b. How do you feel about continuing to vape in the longer term (e.g. months or years)?
16. “What do you think is good and bad about vaping nicotine in the long term”

##### After treatment

17. “Will you consider continuing vape nicotine?

a. If no, what would stop you from continuing to vape nicotine?
b. If you have used other methods to stop smoking in the past, how does vaping compare?
18. “Do you think you would recommend vaping to stop smoking cigarettes to other people? Why or why not?”

#### Analysis

All interviews will be recorded and transcribed. The interview transcripts will be coded manually or may be uploaded into a computer aided qualitative data analysis software (CAQDAS) to assist with analysis.

The content of the interviews will be analysed descriptively, focusing on understanding the patient’s experience with VNPs throughout the study and any major themes identified.

#### Reporting and publication

Data confidentiality, integrity and security will be as per the main protocol.

Results of this study will be used in their own right and in conjunction will data collected from the main study. The investigators plan to discuss/publish the study findings of this research component in publicly available reports, journals, at relevant conferences and other appropriate public forums. Information will be provided in such a way that the participant cannot be identified. No personal details will be released in any publication.

#### Confidentiality

Permission will be sought from participants to audio-record the interview for transcription and analysis purposes. During this process, all identifying information will be removed. Quotes from the discussion may be used in written reports without any distinguishing details included.

